# Estimating the presymptomatic transmission of COVID19 using incubation period and serial interval data

**DOI:** 10.1101/2020.04.02.20051318

**Authors:** Weituo Zhang

## Abstract

We estimated the fraction and timing of presymptomatic transmissions of COVID19 with mathematical models combining the available data of the incubation period and serial interval. We found that up to 79.7% transmissions could be presymptomatic among the imported cases in China outside Wuhan. The average timing of presymptomatic transmissions is 3.8 days (SD = 6.1) before the symptom onset, which is much earlier than previously assumed.

## Introduction

The pandemic of COVID19 is ongoing. Till April 2 2020, COVID19 has caused more than 965 thousand infections and 49 thousand deaths world widely. Current evidences indicate COVID19 can be transmitted presymptomatically [1-2]. But several crucial features of COVID19 transmission behavior remain uncertain: what was the proportion of transmissions happened before symptom onset, and when did the infected persons become infectious? The answers to these questions may significantly impact the disease control strategy [3].

We tried to answer these questions by estimating the infection time distribution of COVID19. The infection time is the time of the secondary case infection relative to the symptom onset of the index case. The fraction and timing of presymptomatic transmissions can be straightforwardly obtained from the distribution of infection time.

As illustrated in Figure 1, the infection time is related to the other two important epidemic features as

**Figure 1:**
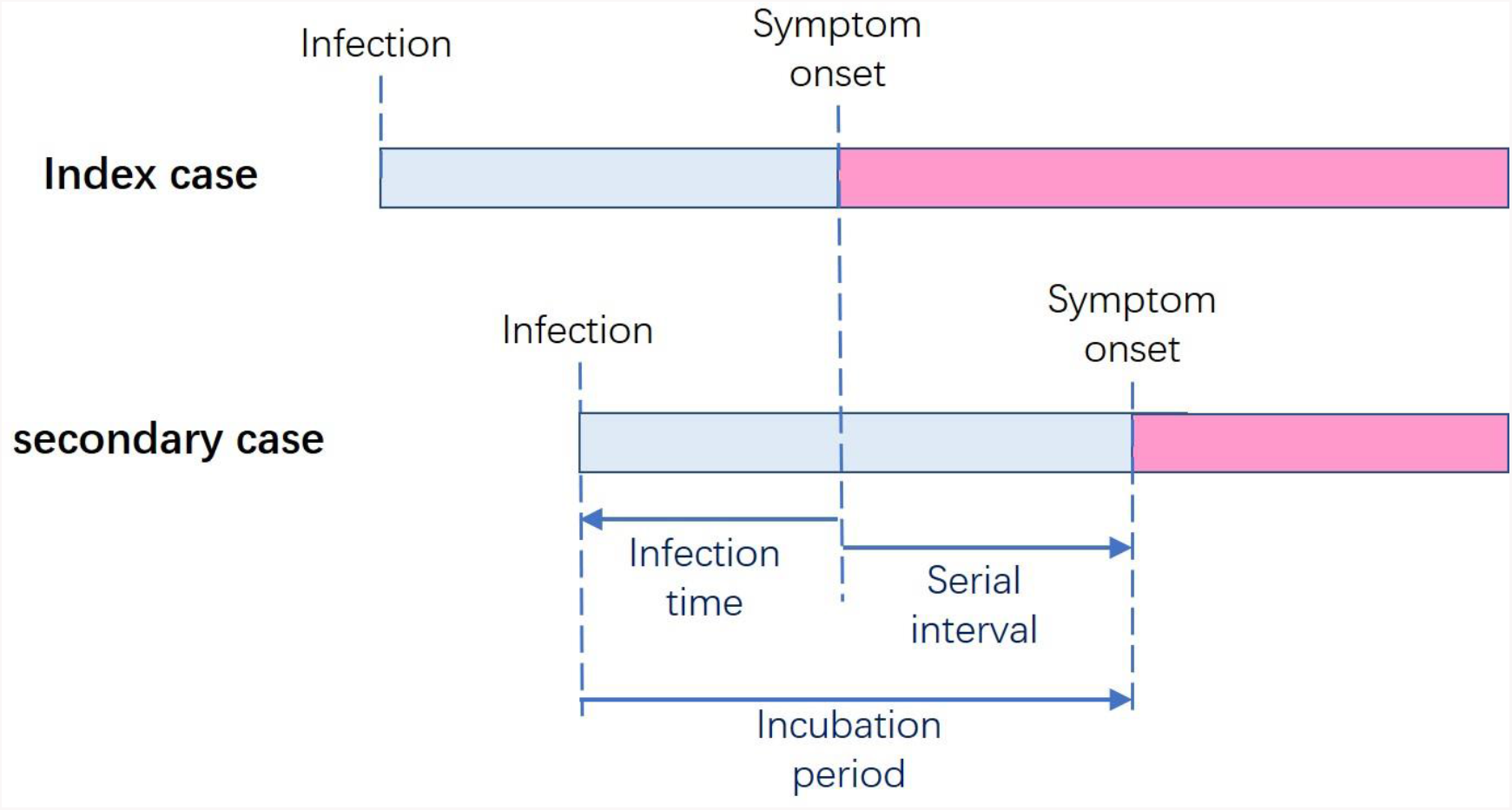
**Illustration of the relationship among the infection time, incubation period and serial interval. The infection time is defined as “date of the secondary case infection - date of the index case symptom onset”.**

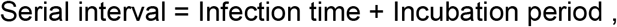

where negative value of infection time stands for the presymptomatic transmission. Given this relationship, the infection time distribution could be estimated using statistical methods combining the up to date data of the incubation period and the serial interval of COVID19.

### Estimate the mean and standard deviation of COVID19 infection time

There have been several reports estimating the incubation period or serial interval of COVID19 [4-8]. Based on these reports and using method of moments (see Appendix 1) we estimated the mean and standard deviation of COVID19 infection time in two scenarios as listed in Table 1: Scenario 1, early transmission in Wuhan, China before Jan. 22, 2020 [4]; Scenario 2, imported transmission in China outside Wuhan after Jan. 20, 2020 [5-6] (the Wuhan lockdown was initialed on Jan. 23, 2020).

**Table 1:** **Estimation of the mean and standard deviation of the infection time of COVID19 in and outside Wuhan based on previous reports of incubation period and serial interval. Following each reference is the time range of data included in the reference.**

|  | Scenario 1<br>Early Transmission in Wuhan |  | Scenario 2<br>Imported Cases Outside Wuhan |  |
| --- | --- | --- | --- | --- |
| | Mean ± Sd<br>(days) | Reference | Mean ± Sd<br>(days) | Reference |
| Incubation Period | $5.2 \pm 2.1^*$ | Li et al. [4]<br>(before Jan. 20,2020) | $6.4 \pm 2.3$ | Baker et al. [5]<br>(Jan. 20-28, 2020) |
| Serial Interval | $7.5 \pm 3.4$ | Li et al. [4]<br>(before Jan. 20,2020) | $3.96 \pm 4.75$ | Du et al. [6]<br>(Jan. 21-Feb. 8, 2020) |
| Infection time | $2.3 \pm 2.7$ | Our estimation | $-2.4 \pm 4.2$ | Our estimation |

\* This standard deviation of incubation period were estimated using gamma distribution.

We estimated that the infection time was 2.3±2.7 days in Scenario 1, and was **-** 2.4±4.2 days in Scenario 2, which implied that the majority of transmissions were post-symptomatic in Scenario 1 but were presymptomatic in Scenario 2. The dramatic change of infection time distribution between these two scenarios may due to effective case isolation and quarantine of people with Wuhan travel history which could significantly reduce transmissions after symptom onset. The method of moments estimations do not rely on any specific assumption of the distribution forms of the incubation period or serial interval.

### Modeling the timing of presymptomatic transmission

We tried to model the timing of COVID19 presymptomatic transmission. We assumed the probability for patients being infectious is given by (more details are provided in Appendix 2)

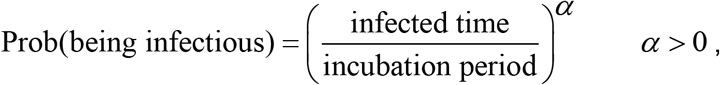

where *α* is a parameter shaping the transmission behavior. As showed in Figure 2, the patients’ probability of being infectious increases from zero at their first day of infection to one at the day of symptom onset. *α* = 1 stands for the assumption that the onsets of infectiousness uniformly distributed in the incubation period, therefore the probability of being infectious increases linearly with time. When *α* > 1, more patients would only become infectious shortly before their symptom onsets, which was previously assumed for COVID19. On the other hand, when *α* < 1, more patients would become infectious shortly after their infections and much earlier than their symptom onsets.

**Figure 2:**
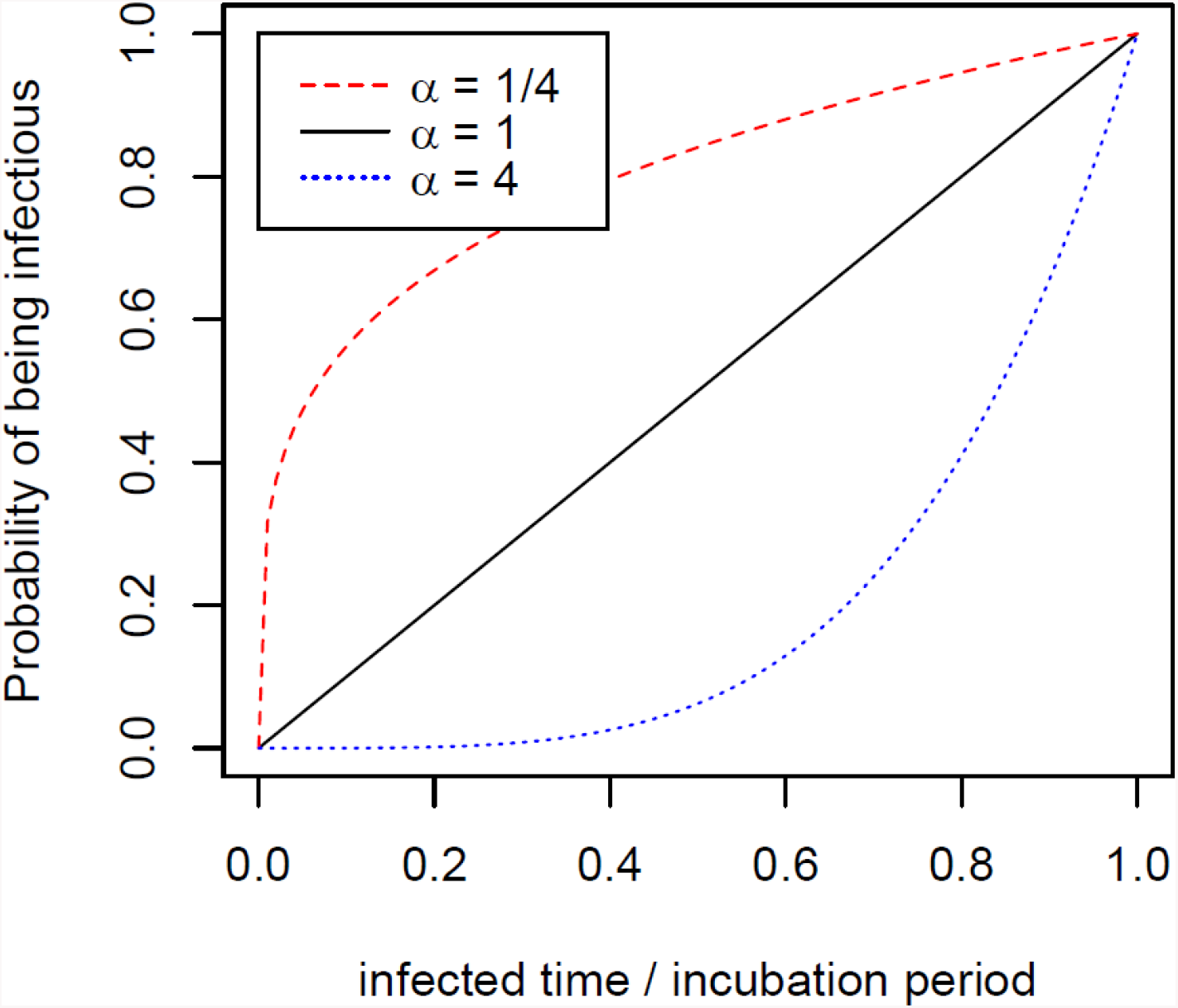
**Presymptomatic transmission behavior model of COVID19 with different *α*. The three curves represent how the probabilities of being infectious increase with time during the incubation period.**

Then serial interval distribution could be calculated using convolution (see Appendix 3).

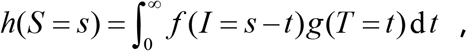

Where *h*(*S*), *f*(*I*), *g*(*T*) are the probability density functions of serial interval, infection and incubation period respectively.

A distinctive feature of our presymptomatic transmission model is to naturally reproduce the “negative serial intervals”, a situation that the symptom onset of the secondary case is earlier than the index case. Widely used statistical models that describing serial interval distributions, including the Gamma, Weibull, log-normal distributions etc., ignore the possibility of negative serial intervals. However, Du et al. reported 12.6% negative serial intervals in the 468 pairs of cases of imported transmissions in China outside Wuhan, which indicated that our model was more proper for COVID19 modeling.

As demonstrated in Figure 3, when *α* = 1/4, 1 and 4, we estimated that the incidence of negative serial intervals were 15.5%, 9.7% and 3.2% respectively among presymptomatic transmissions. Sensitivity analysis using various incubation period distributions was given in Appendix Table S1. Comparing with the observed and model predicted negative serial incidence, it was more likely that *α* < 1, which contradicted with current assumption of COVID19 transmission behavior.

**Figure 3:**
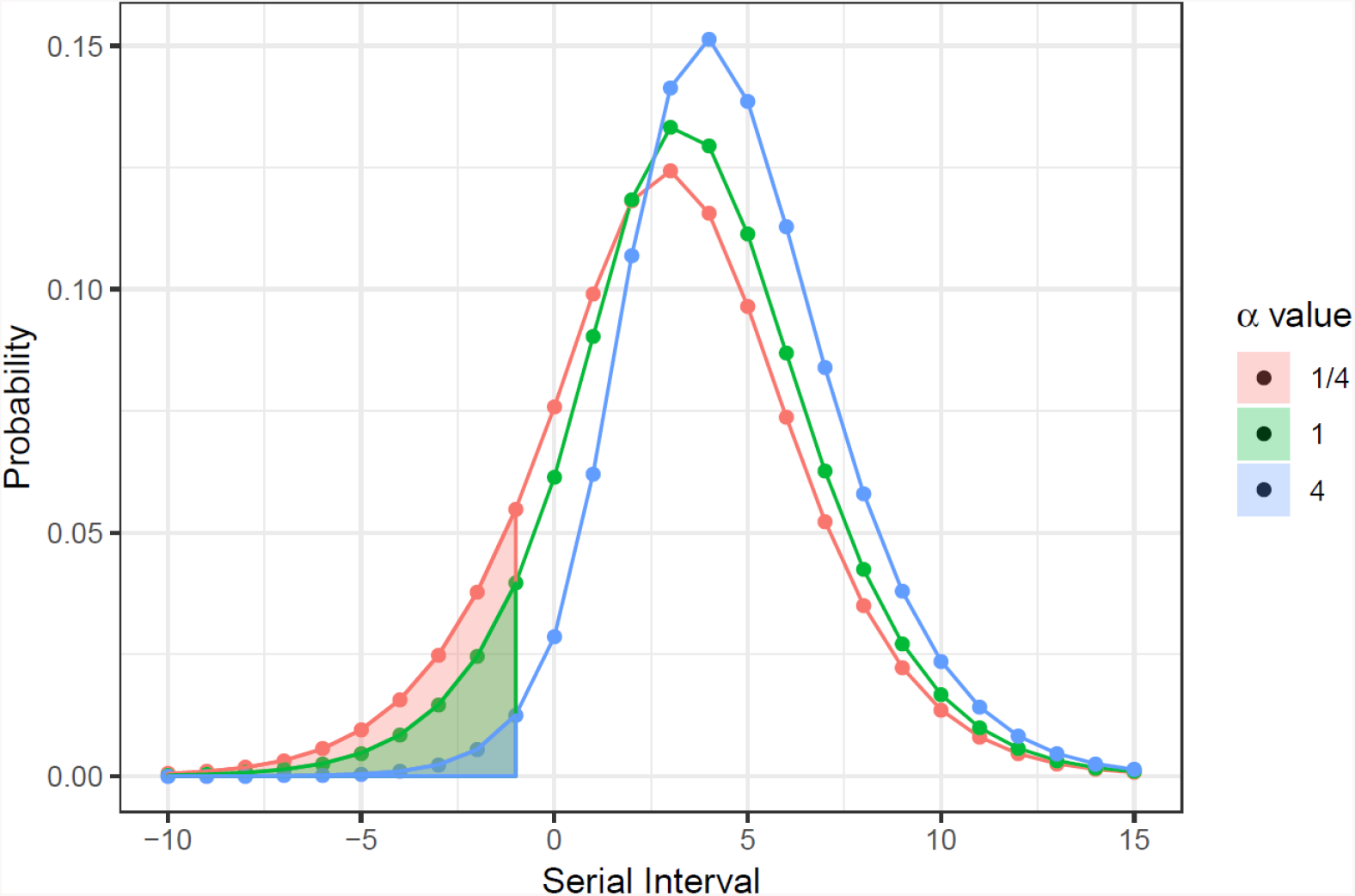
**The serial interval distributions (curves) and the negative serial interval incidences (shadow areas) estimated by the COVID19 presymptomatic transmission behavior model.**

### Estimate the fraction of presymptomatic transmission

Two approaches were applied to estimate the presymptomatic transmission fraction. A raw estimation was based on the mean and standard deviation of infection time estimated previously with normal distribution assumption. The more sophisticated estimation is based on our presymptomatic transmission model and use the maximum convolution likelihood method [9] (see Appendix 4) to estimate the model parameters.

We estimated the distribution of infection time and serial interval in Scenario 2 (as mentioned before) via two approaches as shown in Figure 4. The fraction of presymptomatic transmissions was estimated 79.7% with our model and 67.5% with normal distribution assumption. The *α* parameter was estimated as 0.078, which implied that the COVID19 infected persons tended to become infectious shortly after infection. The average timing of presymptomatic transmission was estimated 3.8 days (SD = 6.1) before symptom onsets.

**Figure 4:**
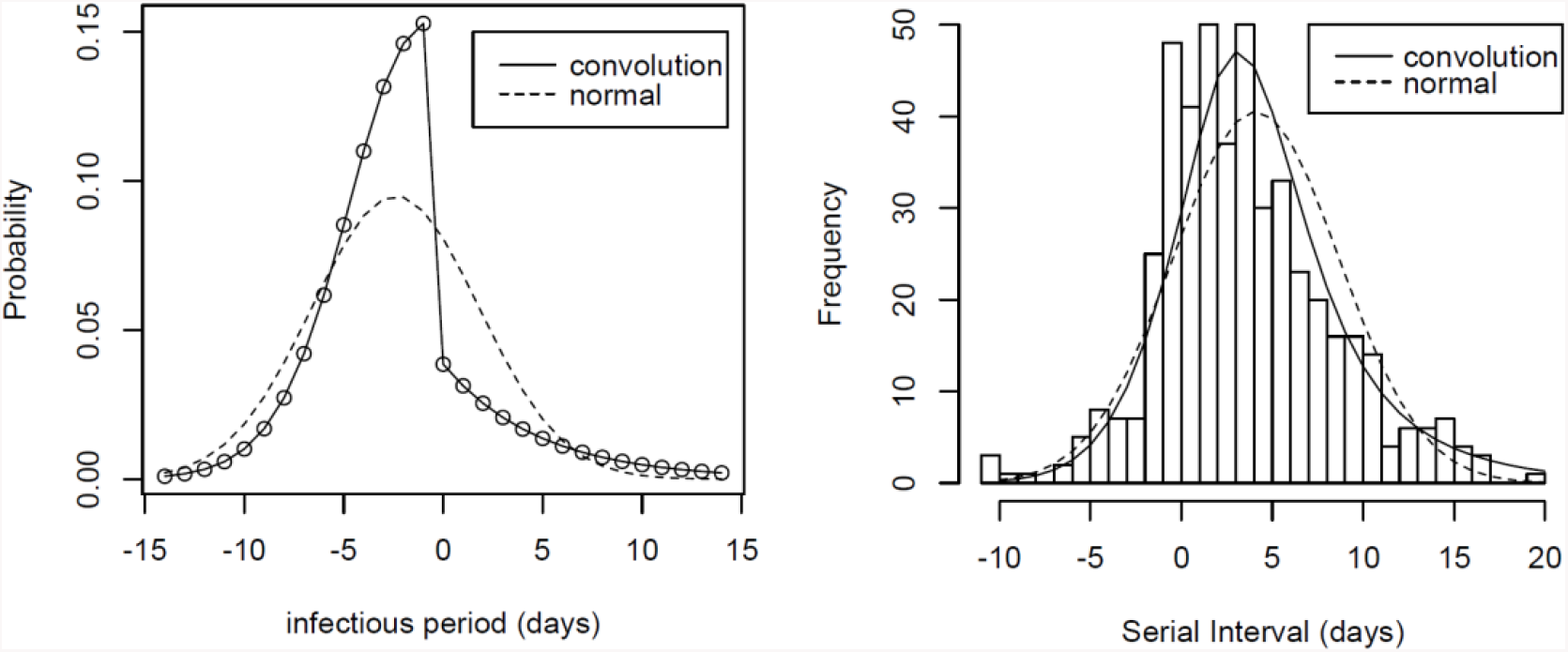
**Estimated infection time distribution (left panel) and serial interval distribution (right panel). The dash lines stand for the estimations with normal distribution assumption, the solid lines stand for our model predictions. The histogram in the right panel is the observed serial interval data.**

Because of the unavailability of data, we could not apply the maximum convolution likelihood estimation on Scenario 1, but the estimation with normal distribution assumption showed 19.7% transmissions were presymptomatic.

## Conclusion and Discussion

In conclusion, we estimated the fraction and timing of COVID19 presymptomatic transmissions. Through several different approaches (method of moments estimation, expected negative serial interval incidence, and maximum convolution likelihood estimation), we achieve largely consistent results: a large portion of COVID19 transmissions may happen presymptomatically. The percentage is roughly 19.7% when there is no effective control, and can go up to 79.7% when control measures effectively reducing transmissions after symptom onsets are performed. The timing of presymptomatic transmission is on average 3.8 days (SD=6.1) before symptom onset. Patients is likely to become infectious in the early stage of their infections instead of just before their symptom onsets.

To our knowledge, our model combining incubation period and serial interval data is a novel approach to estimate the presymptomatic transmissions of infectious disease. The distinctive feature of this approach is that it naturally predict the occurrence of negative serial intervals. The high incidence (12.6%) of negative serial intervals in the observed data of COVID19 (much higher than other diseases like SARS, MERS, etc.) implies that our model is particularly important to correctly understand the COVID19 transmission behavior.

The infection time distribution investigated in this study is closely related to the biologically infectious period of patients during the course of disease. Both provide insights of the transmissibility of COVID19, but from two different aspects. Our findings are largely consistent with the currently available data of viral shedding studies in the close contacts and the confirmed cases [10-11]. However, a difference should be noticed that the infection time distribution is more likely to be affected by the sociological factors and non-pharmaceutical interventions performed by local public health authorities.

Our findings should be interpreted with caution. The epidemic data of COVID19 patients that describing the time and tracing details is very limited. Therefore both estimations of the incubation period and the serial interval of COVID19 have considerable uncertainty. Important biases in our data source include the detection bias(i.e. cases with severe symptoms are more likely to be detected), and reporting bias(i.e. cases with clearly reported tracing details are more likely from areas with plenty of public health resources, and cannot represent the situation in those poorly controlled areas).

Another important concern of COVID19 is the transmissions induced by the “true asymptomatic carrier”, which refers to the infected person who keep asymptomatic during the entire course of disease [12]. Our study is based on data of confirmed cases which did not include the “true asymptomatic carrier”. However either presymptomatic transmission or asymptomatic carrier suggest that the isolations based on symptom surveillance alone may be not enough. Aggressive testing, isolation of close contacts and social distancing to prevent presymptomatic or asymptomatic transmissions are crucial to combat COVID19.

## Data Availability

All data and code to reproduce this paper are available online

https://github.com/witjump/COVID19-presymptomatic-transmission

## Acknowledgement

We appreciate Zhanwei Du et al. collected and cleaned the serial interval data of confirmed cases reported in China, and made the data open for public.

## Appendix Mathematical methods

### 1. Method of moments estimation of infection time

We have

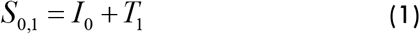

Where *I*_0_ is the infection time of index case, *T*_1_ is the incubation period of secondary case, and *S*_0,1_ is the serial interval. All these three are random variables. It is reasonable to assume *I*_0_ and *T*_1_ are independent. Therefore, we have following equations:

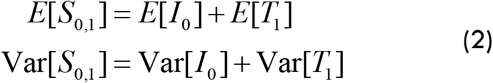

Hence we can estimate the mean and standard deviation of infection time *I*_0_ by solving Eq. (2). Note that, the method of moments estimation does not rely on any assumption of specific distribution forms of the *I*_0_, *T*_1_ and *S*_0,1_, so the conclusion should be robust. But the estimation of infection time can be affected by the bias in previous estimation of incubation period and serial interval.

### 2. Presymptomatic transmission model

For presymptomatic transmission, the infection time of an index case *I*_0_ is restricted by its own incubation period *T*_0_ by *I*_0_ > −*T*_0_. So we model *F*_*i*_ (*I*_*i*_ | *T*_*i*_), the cumulative probability for individual *i* becoming infectious given condition of its incubation period *T*_*i*_, with formula:

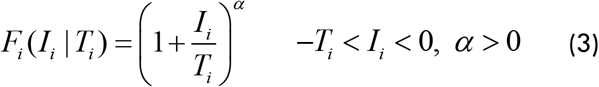

In this model, the probability of infectiousness for an index case increases monotonically from 0 at the time of initial infection (*I*_*i*_ = −*T*_*i*_), to 1 at the time of symptom onset (*I*_*i*_ = 0).

To better understand this infectiousness behavior, let us see a specific case: if an index case develops infectiousness with equal probability at any time during its incubation period, then its probability of being infectious should increase linearly with time. That is the case when *α* = 1. If *α* > 1, the index case is more likely to develop infectiousness shortly before symptom onset, which is the commonly assumed behavior of COVID19 in previous literature. On the contrary, if*α* < 1, the index case is more likely to develop infectiousness shortly after being infected despite the symptom onset might be much later.

The population averaged infection time *f* (*I*) is given by

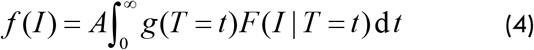

where A is a normalization coefficient. The following Fig. S1 (A) demonstrates the cumulative probability of infectiousness with variety of *α* parameters. The x-axis of Fig. S1(A) is the “infected time over incubation period” (i.e.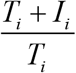, which is an alternative representation of *F*_*i*_ (*I*_*i*_ | *T*_*i*_). Fig. S1(B) demonstrates the population averaged infection time *f* (*I*).

**Figure S1:**
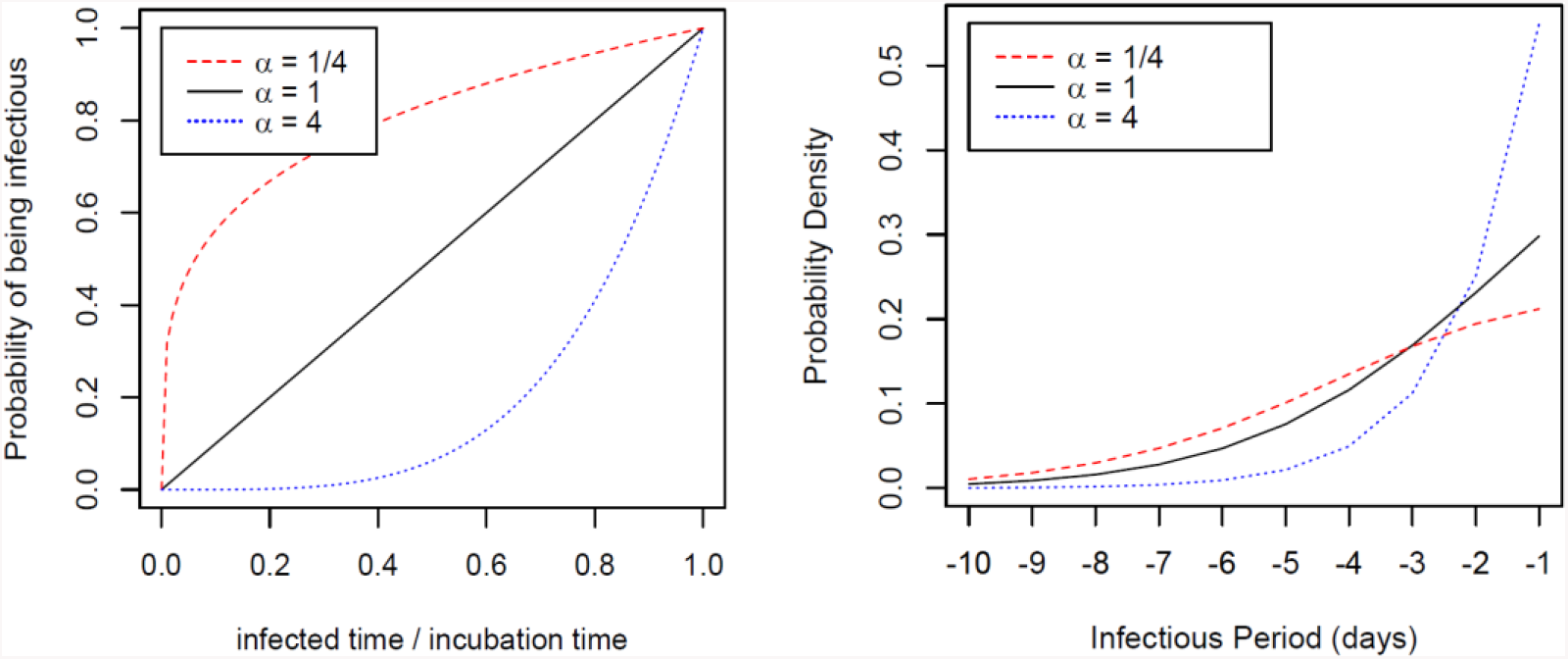
**The individual (left) and population averaged (right) presymptomatic transmission model with different *α*.**

### Part 3. Estimating the serial interval distribution and the incidence of negative serial intervals

Denote the distribution density functions of infection time (population averaged), incubation period and serial interval by *f* (*I*), *g*(*T*), and *h*(*S*) respectively. According to equation (1), we have

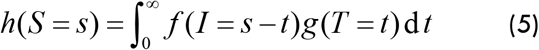

or simply *h* = *f* * *g* with convolution notation. The serial interval distribution *h*(*S*) is calculated using Eq. (5). The expected negative serial fraction is estimated by

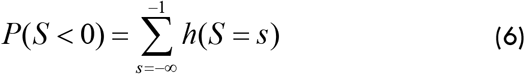

Here we use the discrete sum instead of integration in order to match the empirical data in which the serial time is discrete in days. Besides, we think the negative serial interval less than one day is not observable in realistic setting.

### Part 4: Maximum convolution likelihood estimation

Now we use the empirical serial interval data of COVID19 in China to estimate the infection time distribution.

We construct the whole transmission model with both presymptomatic and post-symptomatic transmission. We use negative binomial distribution to model the infection after symptom onset. Hence the combined infection time distribution is given by

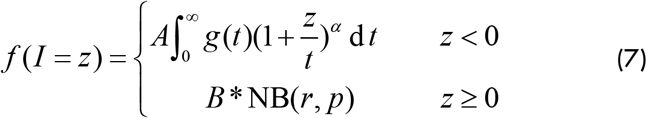

with *α, r, p* and *A* / *B* as undetermined parameters.

Combining Eq. (3),(7), we use maximum likelihood method to estimate the parameters in (7) by maximizing the log-likelihood function

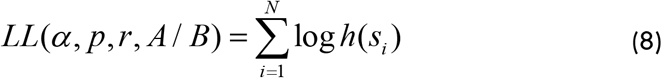

where the *s*_*i*_ (*i* = 1…*N*) are the serial intervals of imported COVID19 cases obtained from China outside Hubei.

**Table S1:** **Sensitivity analysis of the negative serial interval incidence estimation using different incubation period distribution assumptions according to Baker’s report.**

| | $\alpha=1/4$ | $\alpha=1/2$ | $\alpha=1$ | $\alpha=2$ | $\alpha=4$ |
| --- | --- | --- | --- | --- | --- |
| Gamma distribution | 15.5% | 13.2% | 9.7% | 5.5% | 2.2% |
| Weibull distribution | 13.5% | 11.4% | 8.3% | 4.6% | 1.9% |
| Log-normal distribution | 19.0% | 16.5% | 12.5% | 7.8% | 3.5% |

